# Evaluating Polygenic Score Transferability for Lipid Traits in Underrepresented Populations: Evidence from Samoan Cohorts

**DOI:** 10.64898/2026.06.26.26356725

**Authors:** Toni-Ann J. Yapp, Mohanraj Krishnan, Shuwei Liu, Samantha L. Manna, Hong Cheng, Take Naseri, Muagututu’a Sefuiva Reupena, Satupa’itea Viali, John Tuitele, Ranjan Deka, Nicola L. Hawley, Stephen T. McGarvey, Daniel E. Weeks, Ryan L. Minster, Jenna C. Carlson

**Author notes:** **Correspondence:** Toni-Ann J. Yapp, Department of Human Genetics, University of Pittsburgh, 130 De Soto Street, Pittsburgh, PA 15261.

## Abstract

Dyslipidemia is a significant risk factor for cardiovascular disease (CVD), the leading cause of death in Samoa.^1^ Polygenic scores (PGS) for lipid traits offer promise for improved CVD risk prediction,^4,5^ yet their performance in Pacific Islander populations — comprising only 0.002% of GWAS participants as of 2024^3^ — remains unknown. We evaluated the transferability of multi-ancestry PGS for LDL cholesterol (LDL-C), HDL cholesterol (HDL-C), triglycerides (TG), and total cholesterol (TC) in 4,342 Samoan adults across five cohorts spanning 1990–2010. PGS from Graham et al.^8^ and Kanoni et al.^9^ multi-ancestry meta-analyses were harmonized with genome-wide imputed genotypes using a Samoan-specific reference panel^21^ and performance was assessed via incremental R² from linear mixed models with bootstrapped confidence intervals.^25^ HDL-C showed the highest performance (incremental R² 5.0–15.0%), followed by TC (5.0–10.7%), LDL-C (5.7–8.6%), and TG (3.5–7.0%). Critically, meaningful LDL-C performance was achieved only with the genome-wide PRS-CS score (99.6–99.7% variant matching), while a curated pruning-and-thresholding score achieved ∼9% matching and near-zero performance. These findings establish the first systematic lipid PGS benchmarks in Samoans, demonstrating meaningful transferability when genome-wide variant coverage is ensured, and highlight variant harmonization as a critical precondition for PGS deployment in underrepresented populations.

## INTRODUCTION

Dyslipidemia is a major modifiable risk factor for CVD, which is the leading cause of death in Samoa, accounting for 34% of deaths.^1^ PGS aggregate genetic effects across many variants to predict individual-level predisposition to complex traits and have demonstrated utility in risk stratification for lipid-related cardiovascular conditions.^4,5^ However, PGS performance degrades substantially in non-European populations due to differences in allele frequencies, linkage disequilibrium patterns, and underrepresentation in imputation reference panels, such that clinical utility has largely been demonstrated only in populations of European ancestry.^6,7^

Pacific Islanders are among the most underrepresented groups in genomic research, comprising only 0.002% of GWAS participants as of 2024,^3^ yet face disproportionately high burdens of CVD and metabolic disease. No systematic evaluation of lipid PGS transferability has been reported for any Pacific Islander population. Graham et al.^8^ and Kanoni et al.^9^ generated multi-ancestry PGS for all four major lipid traits from approximately 1.65 million individuals across diverse ancestries, representing the most promising existing tools for cross-ancestry prediction. The Samoan population, with well-characterized cohorts spanning two decades and imputed using a Samoan-specific reference panel,^21^ provides an ideal setting to benchmark PGS transferability in this underserved group. Here we report the first systematic lipid PGS benchmarks in Samoans, with particular attention to the role of variant harmonization in determining apparent transferability.

## MATERIALS AND METHODS

### Study Population and Phenotype Data

Five cohorts of Samoan adults were recruited from Samoa and American Samoa between 1990 and 2010. After restricting to participants with non-missing fasting lipid and genotype data, 4,342 participants were included. Detailed sampling, recruitment, and phenotyping procedures have been previously described.^13–18^ Serum lipid levels (TC, HDL-C, LDL-C, TG) were derived from fasting whole blood samples; LDL-C was estimated using the Friedewald equation^19^ and was missing for individuals with TG ≥ 400 mg/dL. Individuals reporting heart disease medication use were excluded.^11^ All participants provided written informed consent in the Samoan language under protocols approved by relevant Institutional Review Boards.

### Genotyping, Quality Control, and Imputation

The 2010 Samoa cohort was genotyped on Affymetrix 6.0 arrays; earlier cohorts on Illumina Global Screening Array-24 v3.0 BeadChips with custom content. Quality control followed Laurie et al.^20^ Genome-wide genotype imputation was performed separately by genotyping platform using a Samoan-specific reference panel;^21^ variants with imputation r² > 0.3 were retained.

### Polygenic Score Implementation

We applied PGS for LDL-C (Graham et al.;^8^ PGS000888, PRS-CS, ∼1.24 million variants), HDL-C (Kanoni et al.;^9^ PGS002781), TG (PGS002784), and TC (PGS002783). Variant weights were downloaded from the PGS Catalog,^22^ lifted over from GRCh37 to GRCh38 using UCSC liftOver,^23^ and harmonized with imputed genotype dosages. Individual-level PGS were computed by summing products of dosage and variant weight. We selected PGS000888 for LDL-C after initial analyses using the alternative pruning-and-thresholding score (PGS000889; 9,009 variants) revealed only ∼9% variant matching in Samoan imputed data, yielding near-zero predictive performance; PGS000888 achieved 99.6–99.7% matching.

### Statistical Analysis

Triglyceride values were natural log-transformed and all traits were inverse-normalized. PGS performance was evaluated using linear mixed models for each trait and cohort, with PGS as the primary predictor and age, age², sex, age×sex, age²×sex, and the first three principal components of ancestry as covariates; kinship was modeled as a random effect via an empirical kinship matrix (lmekin, coxme R package v2.2-18).^24^ Incremental R² (difference in adjusted R² between full and covariate-only models) and partial R² were calculated with 95% confidence intervals from 1,000 bootstrap resamples.^25^ All analyses were performed in R v4.3.1.

## RESULTS

### Study Population

The five Samoan cohorts included 4,342 participants with complete lipid and genotype data (Table 1). The largest was the 2010 Samoa cohort (n = 2,851); earlier cohorts ranged from 169 to 529 participants. Cohort-level lipid distributions showed variation across time periods and geographic locations, particularly for HDL-C and TG, consistent with documented environmental and lifestyle transitions during this period.^13,26^

**Table 1.** Participant characteristics by cohort.

| Cohort | N | %F | Age, mean<br>(SD) | TC, mean<br>(SD) | HDL-C,<br>mean (SD) | LDL-C,<br>mean (SD) | TG, mean<br>(SD) |
| --- | --- | --- | --- | --- | --- | --- | --- |
| 2010 Samoa | 2,851 | 59.8 | 45.1 (11.2) | 199.7 (37.4) | 45.2 (11.0) | 130.2 (33.4) | 126.1 (97.8) |
| 1995 Samoa | 383 | 52.8 | 39.2 (8.6) | 199.4 (36.9) | 43.9 (10.6) | 139.1 (33.2) | 85.0 (76.4) |
| 1994 Am.<br>Samoa | 169 | 57.4 | 40.0 (9.1) | 197.3 (30.2) | 33.6 (8.2) | 136.0 (33.8) | 139.9 (105.8) |
| 2003 Samoa | 410 | 51.0 | 41.8 (16.9) | 197.6 (37.5) | 46.8 (10.8) | 128.8 (34.7) | 112.0 (74.2) |
| 2002 Am.<br>Samoa | 529 | 58.0 | 43.1 (16.1) | 188.0 (38.2) | 40.6 (8.8) | 118.2 (34.2) | 157.3 (135.0) |

TC = total cholesterol; HDL-C = HDL cholesterol; LDL-C = LDL cholesterol; TG = triglycerides; SD = standard deviation; %F = percent female. Values are mean (SD). LDL-C estimated using the Friedewald equation;<sup>19</sup> missing for individuals with TG ≥ 400 mg/dL.

### PGS Variant Harmonization

Variant matching rates were high across all PGS and both genotyping platforms (96–100%; Table 2). The genome-wide LDL-C (PGS000888) and HDL-C (PGS002781) scores achieved 99.6–99.7% matching. TC and TG scores showed lower matching rates (96.1–96.7%) and substantially higher proportions of monomorphic variants in Samoans (12.8–27.6% for TC and TG versus 1.6–4.4% for HDL-C and LDL-C). When we evaluated the curated LDL-C pruning-and-thresholding score (PGS000889), the variant matching rate was only ∼9%, yielding near-zero predictive performance across all cohorts; all reported LDL-C results therefore use PGS000888.

**Table 2.** PGS variant harmonization by trait and genotyping platform.

| Trait | Platform | Total variants | Matched | Match rate | Monomorphic | Mono rate | Mean R<sup>2</sup> |
| --- | --- | --- | --- | --- | --- | --- | --- |
| HDL-C | Affymetrix 6.0 | 1,238,756 | 1,234,496 | 99.7% | 53,796 | 4.4% | 0.937 |
| LDL-C | Affymetrix 6.0 | 1,238,615 | 1,234,493 | 99.7% | 53,796 | 4.4% | 0.937 |
| TC | Affymetrix 6.0 | 10,699 | 10,279 | 96.1% | 1,767 | 17.2% | 0.660 |
| TG | Affymetrix 6.0 | 30,071 | 29,063 | 96.6% | 8,027 | 27.6% | 0.550 |
| HDL-C | Illumina GSA | 1,238,756 | 1,233,296 | 99.6% | 20,255 | 1.6% | 0.921 |
| LDL-C | Illumina GSA | 1,238,615 | 1,233,293 | 99.6% | 20,254 | 1.6% | 0.921 |
| TC | Illumina GSA | 10,699 | 10,288 | 96.2% | 1,316 | 12.8% | 0.694 |
| TG | Illumina GSA | 30,071 | 29,088 | 96.7% | 5,847 | 20.1% | 0.583 |

Affymetrix platform: 2010 Samoa cohort (n=2,851). Illumina GSA platform: all earlier cohorts. Monomorphic rate: proportion of matched variants with MAF=0 in Samoans. Mean R<sup>2</sup>: mean imputation quality score.

### Polygenic Score Performance

Incremental R² and partial R² estimates for all traits and cohorts are presented in Table 3 and Figure 1. HDL-C showed the strongest overall performance (incremental R²: 5.0–15.0% across cohorts; 2010 Samoa: 12.0%, 95% CI: 9.8–14.1%). TC showed broadly consistent performance (range: 5.0–10.7%; 2010 Samoa: 8.8%, 95% CI: 7.0–10.7%). LDL-C using PGS000888 showed meaningful and consistent performance across all cohorts (range: 5.7–8.6%; 2010 Samoa: 7.5%, 95% CI: 5.7–9.4%). TG showed the lowest performance (range: 3.5–7.0%; 2010 Samoa: 4.8%, 95% CI: 3.3–6.4%). Confidence intervals were widest in the smallest cohort (1994 American Samoa, n = 169). Partial R² estimates closely tracked incremental R² and are presented in full in Table 3.

**Figure 1.**
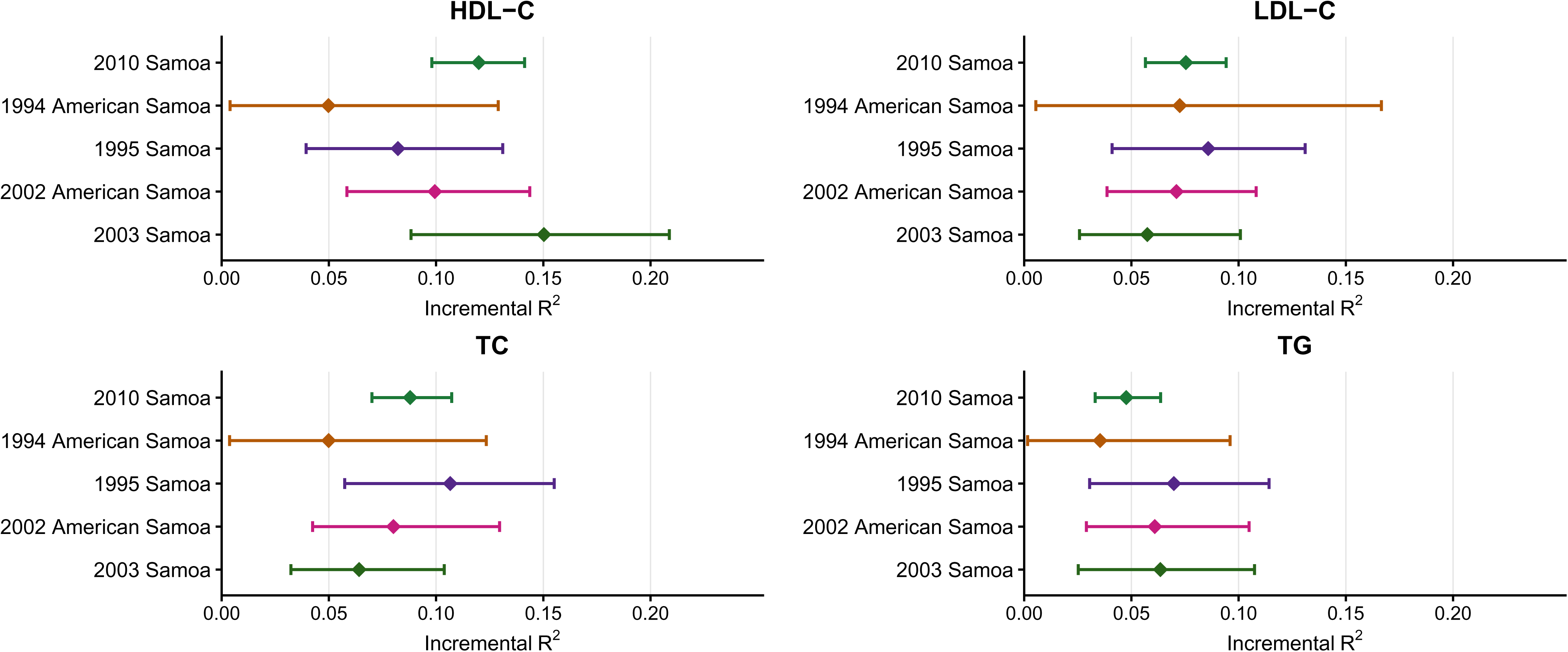
Polygenic Risk Score Performance Across Cohorts and Traits. Incremental R² represents the proportion of phenotypic variance explained by the PGS beyond that explained by age, age², sex, age×sex interactions, and the first three principal components of ancestry. 95% confidence intervals were calculated using bootstrap resampling with 1,000 iterations. WS = Western Samoa (now Samoa); AS = American Samoa.

**Table 3.**
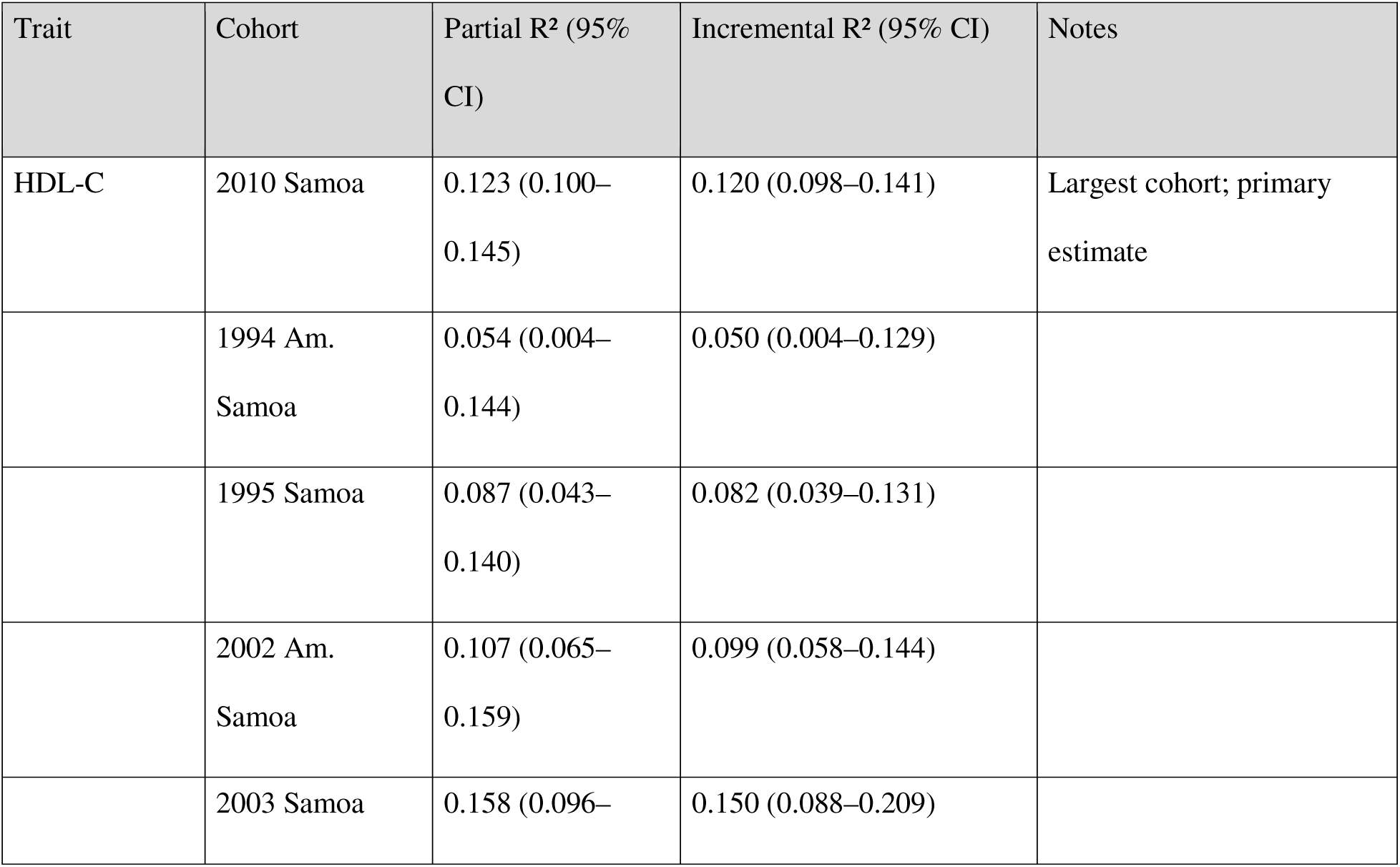
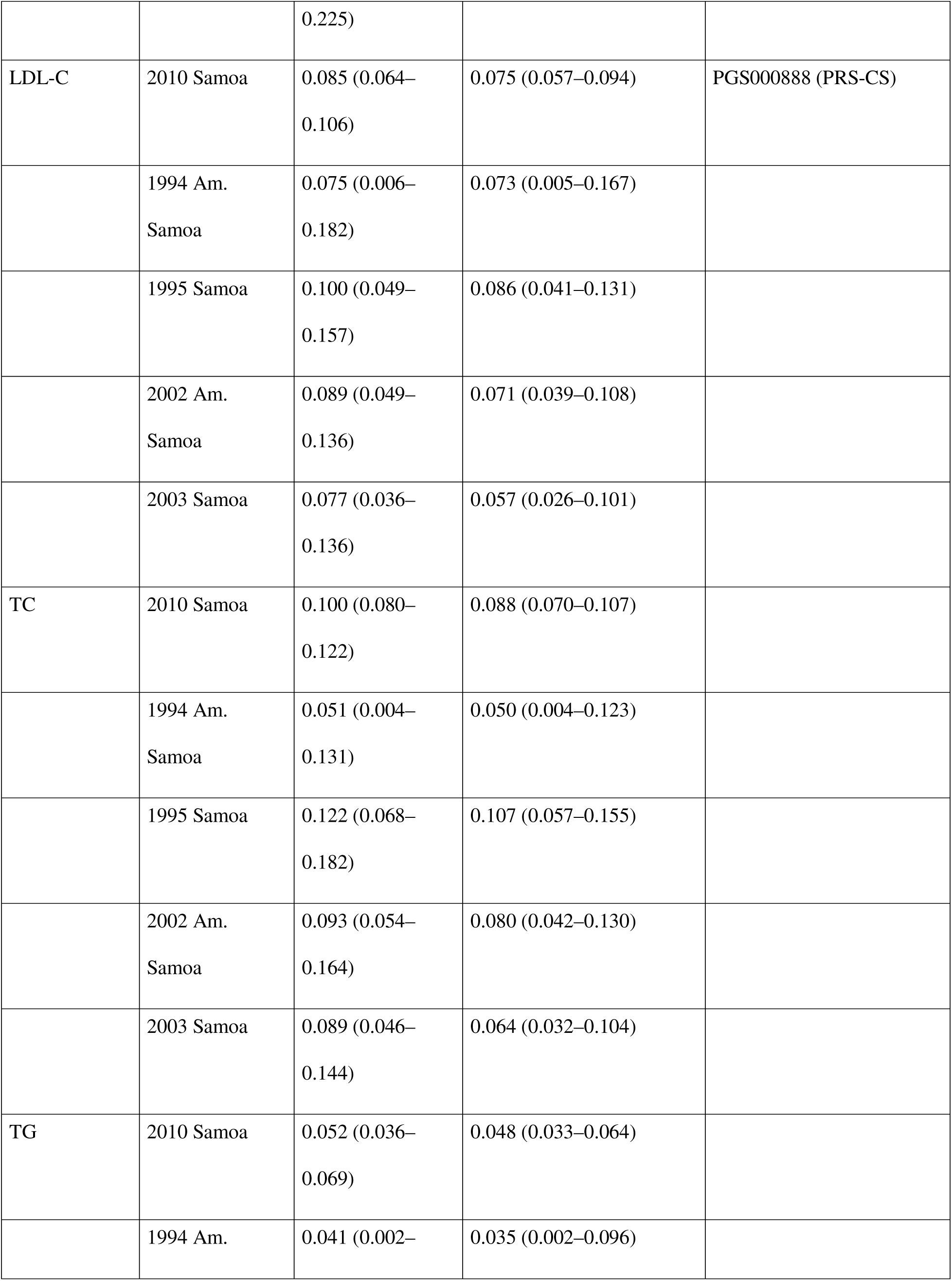
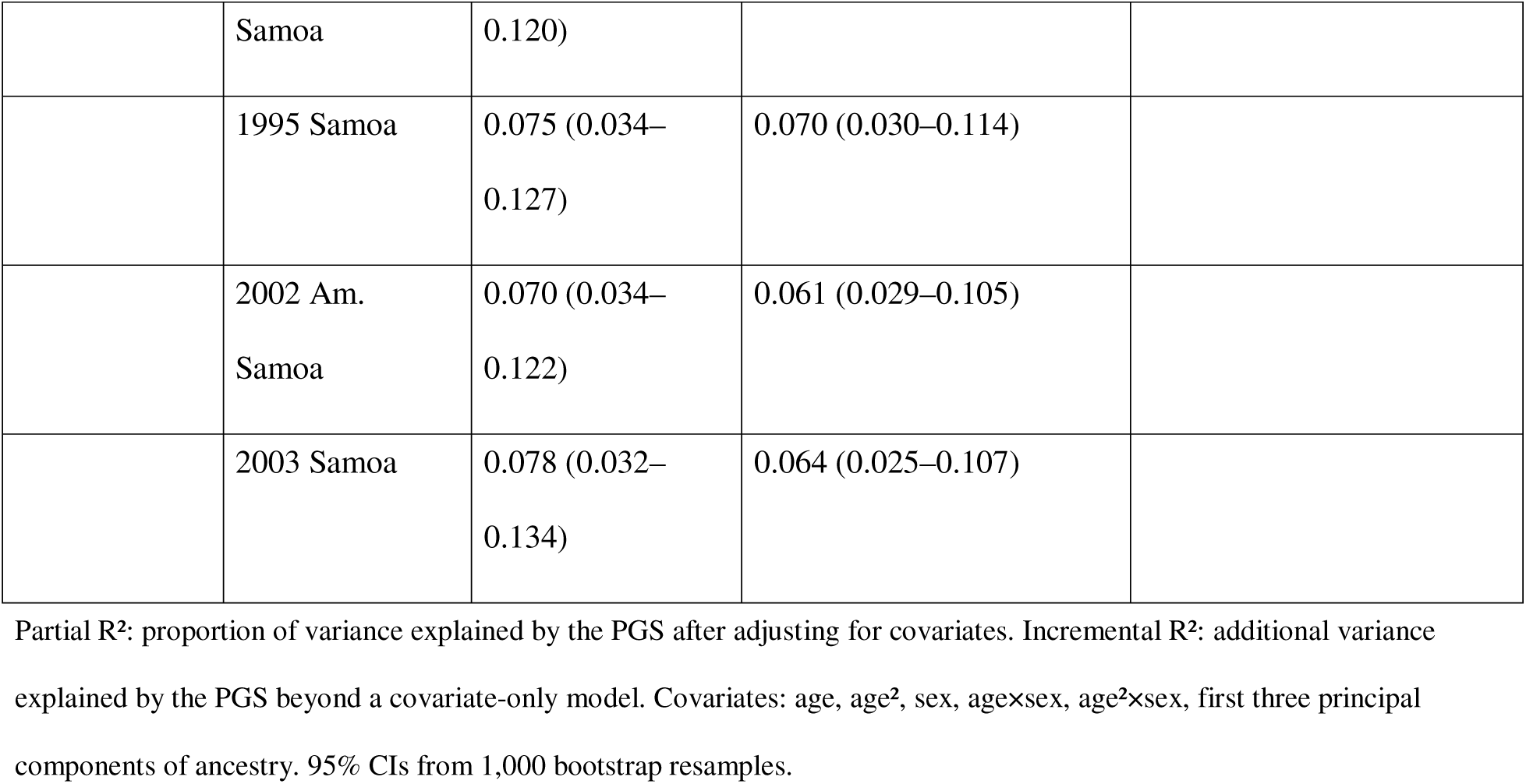
PGS performance for four lipid traits across five Samoan cohorts.

## DISCUSSION

We report the first systematic lipid PGS benchmarks in a Pacific Islander population, evaluating multi-ancestry scores for LDL-C, HDL-C, TG, and TC across 4,342 Samoan adults in five cohorts spanning 20 years. The most consequential finding is the critical dependence of apparent LDL-C transferability on score construction. The curated pruning-and-thresholding score (PGS000889; 9,009 variants) achieved only ∼9% variant matching in Samoan imputed data and near-zero predictive performance, while the genome-wide PRS-CS score (PGS000888; ∼1.24 million variants) achieved 99.6–99.7% matching and meaningful performance (incremental R²: 5.7–8.6%). This provides a direct illustration of a broadly applicable principle: curated small-variant scores may fail entirely in underrepresented populations whose haplotype structure differs from the discovery cohort, even when genome-wide scores from the same GWAS perform adequately. Our results suggest that some published reports of poor LDL-C PGS transferability in non-European populations may reflect score construction choices rather than fundamental differences in genetic architecture, and we recommend that variant matching rates and monomorphic variant proportions be reported as a standard component of any PGS transferability assessment.

Differential performance across traits is consistent with genetic architecture and variant-level characteristics. HDL-C and TC, both highly heritable traits^27,28^ with large-effect loci (*CETP*, *LIPC*, *APOE*) that replicate across ancestries and have been confirmed in Samoan GWAS,^11^ showed the strongest and most consistent performance. TG consistently showed the lowest performance, likely reflecting elevated monomorphic variant rates (20.1–27.6%) and strong environmental modulation: TG is acutely sensitive to dietary and lifestyle factors, and the Samoan cohorts span a period of rapid transition toward modernized diets and activity patterns.^13,26^ The resulting high environmental variance may attenuate the detectable genetic signal.

To contextualize our results, Graham et al.^8^ reported partial R² for LDL-C PGS of approximately 10–11% in European populations, 8–10% in East Asian and South Asian populations, and 5–7% in African populations. Our Samoan LDL-C partial R² estimates (7.5–10.0%) fall within the range reported for East Asian and South Asian populations, suggesting the genome-wide PRS-CS approach achieves broadly comparable transferability in Samoans to other non-European populations in the discovery cohort. Similarly, HDL-C and TC partial R² estimates are broadly consistent with non-European benchmarks from Kanoni et al.^9^ Recent work evaluating PGS in

Native Hawaiians — a population with substantial Polynesian ancestry — found reduced prediction accuracy relative to European benchmarks and highlighted the persistent challenge of limited Pacific Islander representation in discovery GWAS.^29,30^ Our findings are consistent with this picture, while specifically demonstrating that variant-level harmonization decisions are at least as important as population-level genetic architecture in determining observed transferability.

Several limitations merit acknowledgment. Sample sizes, while among the largest available for Pacific Islander genetic studies, remain modest compared to European GWAS, producing wide confidence intervals particularly in smaller cohorts. The temporal span of cohorts (1990–2010) coincides with rapid lifestyle transitions in Samoa and American Samoa;^13,26^ differences in PGS performance across cohorts could reflect gene–environment interactions rather than genetic heterogeneity, and formal interaction analyses represent an important future direction.

Generalizability to other Pacific Islander populations — including Native Hawaiians, Tongans, and Māori — requires dedicated investigation, as these groups are genetically and culturally distinct. Finally, all PGS evaluated here were derived from multi-ancestry studies that did not include Pacific Islanders; dedicated inclusion of Samoan and other Pacific Islander cohorts in future GWAS discovery efforts would likely yield further improvements, particularly for TG, whose lower performance plausibly reflects population-specific genetic architecture not captured by existing reference panels.

## SUPPLEMENTAL INFORMATION

Supplemental Information includes eight figures (Figures S1–S8). Figures S1–S4 show distributions of observed lipid values by cohort. Figures S5–S8 show the relationships between observed lipid values and corresponding PGS scores across cohorts for each trait.

## Data Availability

The 2010 cohort data are available through dbGaP (accession numbers: phs000914.v1.p1 and phs000972.v5.p1). The data from earlier cohorts have not been deposited in a public repository because participants did not give consent for data sharing.

## ACKNOWLEDGMENTS

We thank the Samoan and American Samoan participants of these studies, the local village authorities, and the field workers who contributed over the years. We acknowledge the Samoa Ministry of Health, the American Samoa Department of Health, village councils (*fono*), and traditional leaders (*matai*) for their support and guidance. We thank the Samoan Health Research Committee, the American Samoa Institutional Review Board, and village leaders who facilitated community engagement and participant recruitment. We give particular thanks to research assistants Melania Selu and Vaimoana Lupematasila, who contributed to the 2010 recruitment and continue to assist us in our work in Samoa, and to the many other research assistants, community health workers, and village women’s committees (*komiti tumama*) who supported data collection across all study periods.

Molecular data for the Trans-Omics in Precision Medicine (TOPMed) program was supported by the NHLBI. Genome sequencing for NHLBI TOPMed: Samoan (phs000972.v5.p1) was performed at New York Genome Center Genomics (HHSN268201500016C) and Northwest Genomics Center (HHSN268201100037C). Core support was provided by the TOPMed Informatics Research Center (3R01HL-117626-02S1; contract HHSN268201800002I) and TOPMed Data Coordinating Center (R01HL-120393; U01HL-120393; contract HHSN268201800001I).

## AUTHOR CONTRIBUTIONS

T.J.Y.: Conceptualization, Formal analysis, Methodology, Software, Visualization, Writing – original draft, Writing – review & editing. M.K.: Investigation (GWAS data generation). S.L.: Data curation (WGS pipeline). S.L.M.: Data curation (WGS pipeline), Writing – review & editing. H.C.: Data curation, Investigation. T.N.: Investigation, Resources. M.S.R.: Investigation, Resources. S.V.: Investigation, Resources. J.T.: Investigation, Resources. R.D.: Funding acquisition, Resources. N.L.H.: Conceptualization, Funding acquisition, Investigation, Project administration, Resources. S.T.M.: Conceptualization, Funding acquisition, Investigation, Project administration, Resources. D.E.W.: Resources, Writing – review & editing. R.L.M.: Funding Acquisition, Resources, Writing – review & editing. J.C.C.: Conceptualization, Resources, Supervision, Writing – review & editing.

## WEB RESOURCES

PGS Catalog: https://www.pgscatalog.org/

dbGaP: https://www.ncbi.nlm.nih.gov/gap/ (phs000914.v1.p1; phs000972.v5.p1) Analysis code: [GitHub URL to be added upon acceptance]

## DATA AND CODE AVAILABILITY

The 2010 cohort data are available through dbGaP (phs000914.v1.p1 and phs000972.v5.p1). Data from earlier cohorts are available from the corresponding author upon reasonable request. Analysis code will be made available via GitHub upon acceptance.

## ETHICS APPROVAL STATEMENT

All procedures were in accordance with the ethical standards of relevant institutional and national research committees and the 1964 Helsinki declaration. The study was approved by the IRB of The Miriam Hospital (1994 and 1995 cohorts), the Brown University IRB (2002, 2003, and 2010 cohorts), the University of Pittsburgh IRB, the American Samoa Department of Health IRB, and the Samoa Ministry of Health Health Research Committee.

