## Supplementary material for "Evaluating Polygenic Score Transferability for Lipid Traits in Underrepresented Populations: Evidence from Samoan Cohorts": S1-S8

### Table of Contents


Figure S1. HDL−C: observed levels vs. standardized PGS (raw) 2

Figure S2. LDL−C: observed levels vs. standardized PGS (raw) 3

Figure S3. TC: observed levels vs. standardized PGS (raw) 4

Figure S4. TG: log−transformed levels vs. standardized PGS (raw) 5

Figure S5. HDL−C: covariate−adjusted partial residuals vs. PGS 6

Figure S6. LDL−C: covariate−adjusted partial residuals vs. PGS 7

Figure S7. TC: covariate−adjusted partial residuals vs. PGS 8

Figure S8. TG: covariate−adjusted log(TG) partial residuals vs. PGS 9

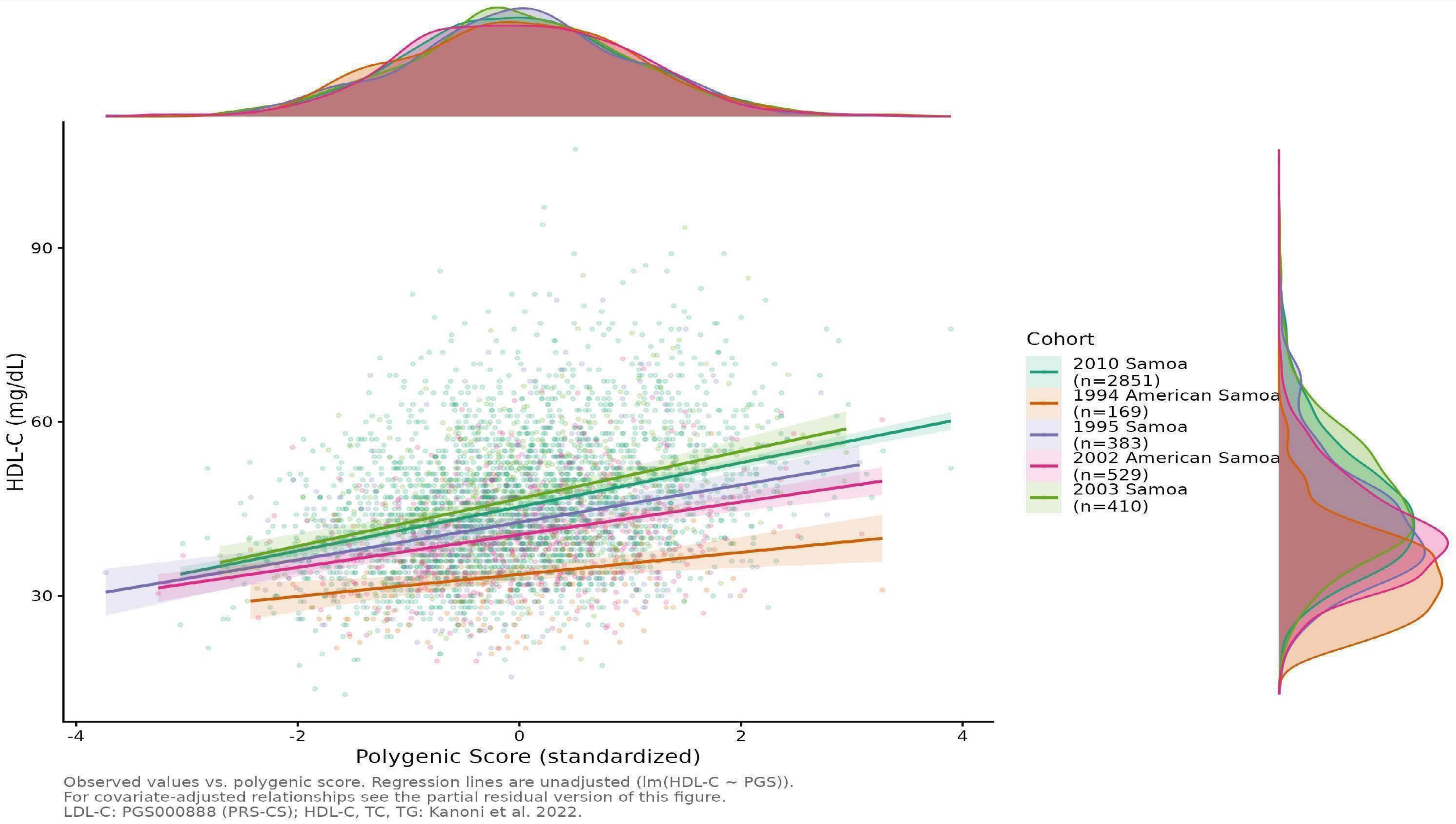


Figure S1. Distribution and polygenic score associations for HDL−C across five Samoan cohorts. Observed HDL−C levels (mg/dL) vs. standardized polygenic score (PGS), with unadjusted linear regression lines and 95% confidence bands per cohort. Marginal density plots show the distribution of PGS (top) and HDL−C levels (mg/dL) (right) by cohort. PGS source: Kanoni et al. (2022).

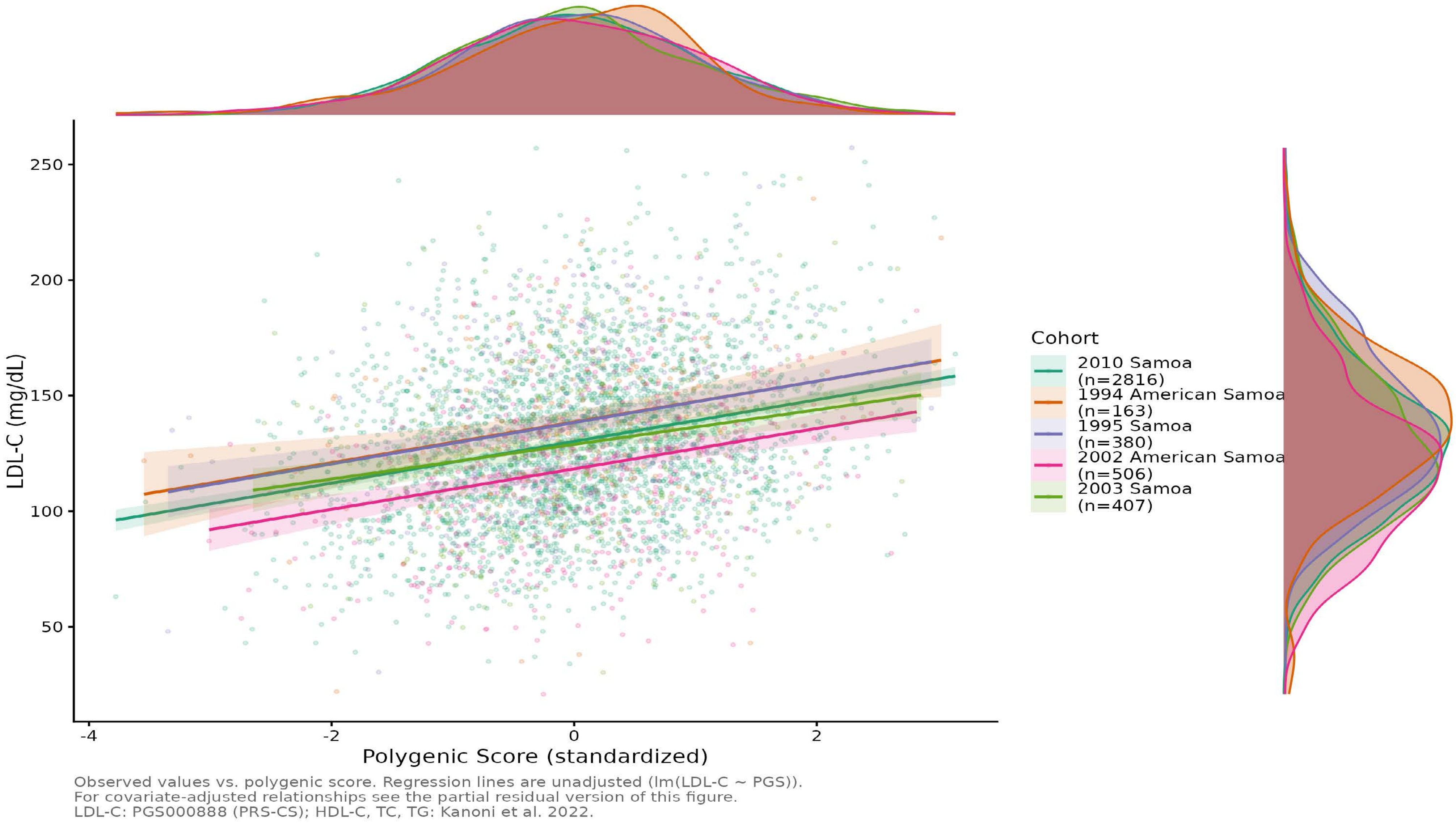


Figure S2. Distribution and polygenic score associations for LDL−C across five Samoan cohorts. Observed LDL−C levels (mg/dL) vs. standardized polygenic score (PGS), with unadjusted linear regression lines and 95% confidence bands per cohort. Marginal density plots show the distribution of PGS (top) and LDL−C levels (mg/dL) (right) by cohort. PGS source: PGS000888, Graham et al. (2021), PRS−CS method.

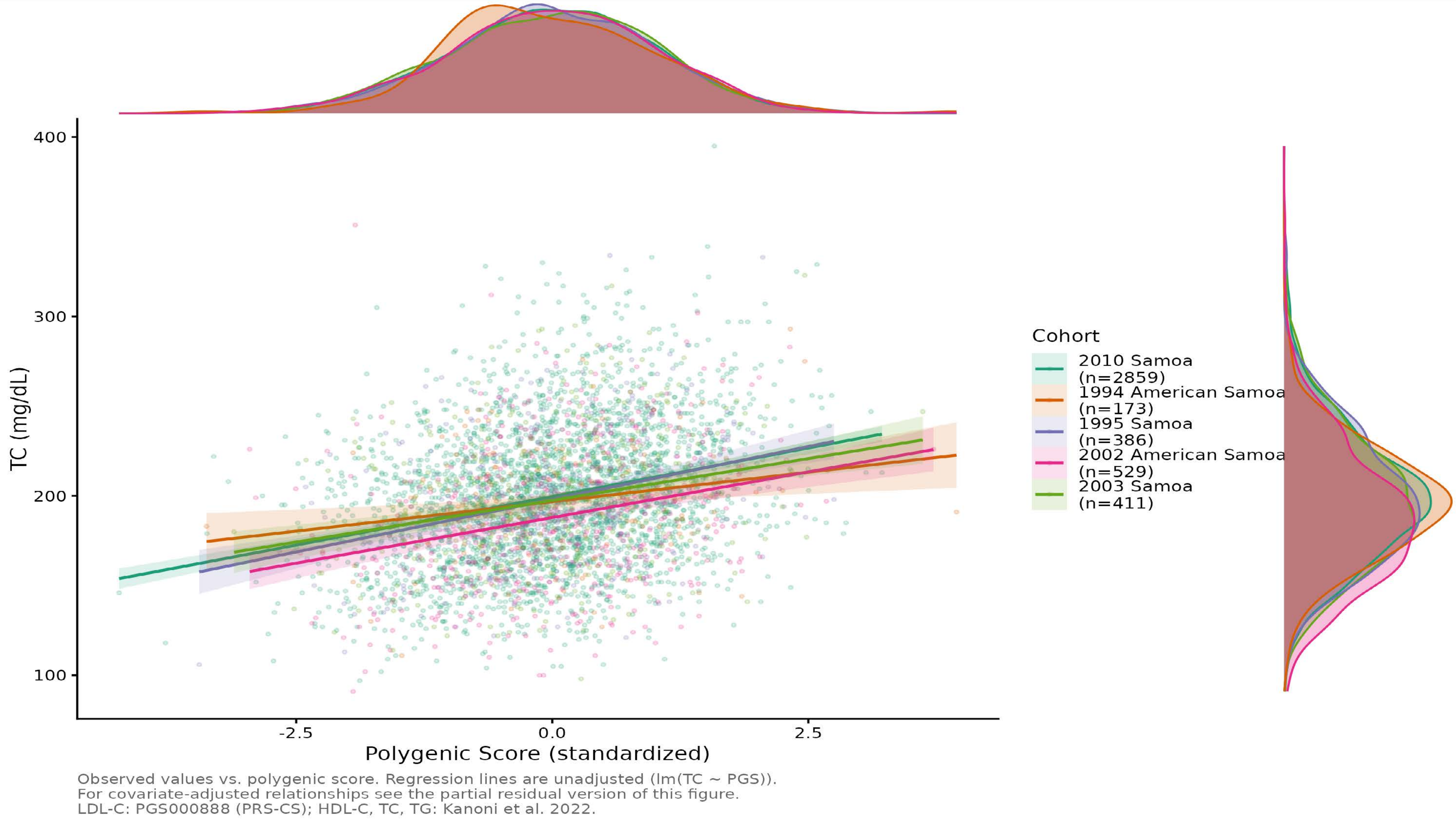


Figure S3. Distribution and polygenic score associations for total cholesterol (TC) across five Samoan cohorts. Observed TC levels (mg/dL) vs. standardized polygenic score (PGS), with unadjusted linear regression lines and 95% confidence bands per cohort. Marginal density plots show the distribution of PGS (top) and TC levels (mg/dL) (right) by cohort. PGS source: Kanoni et al. (2022).

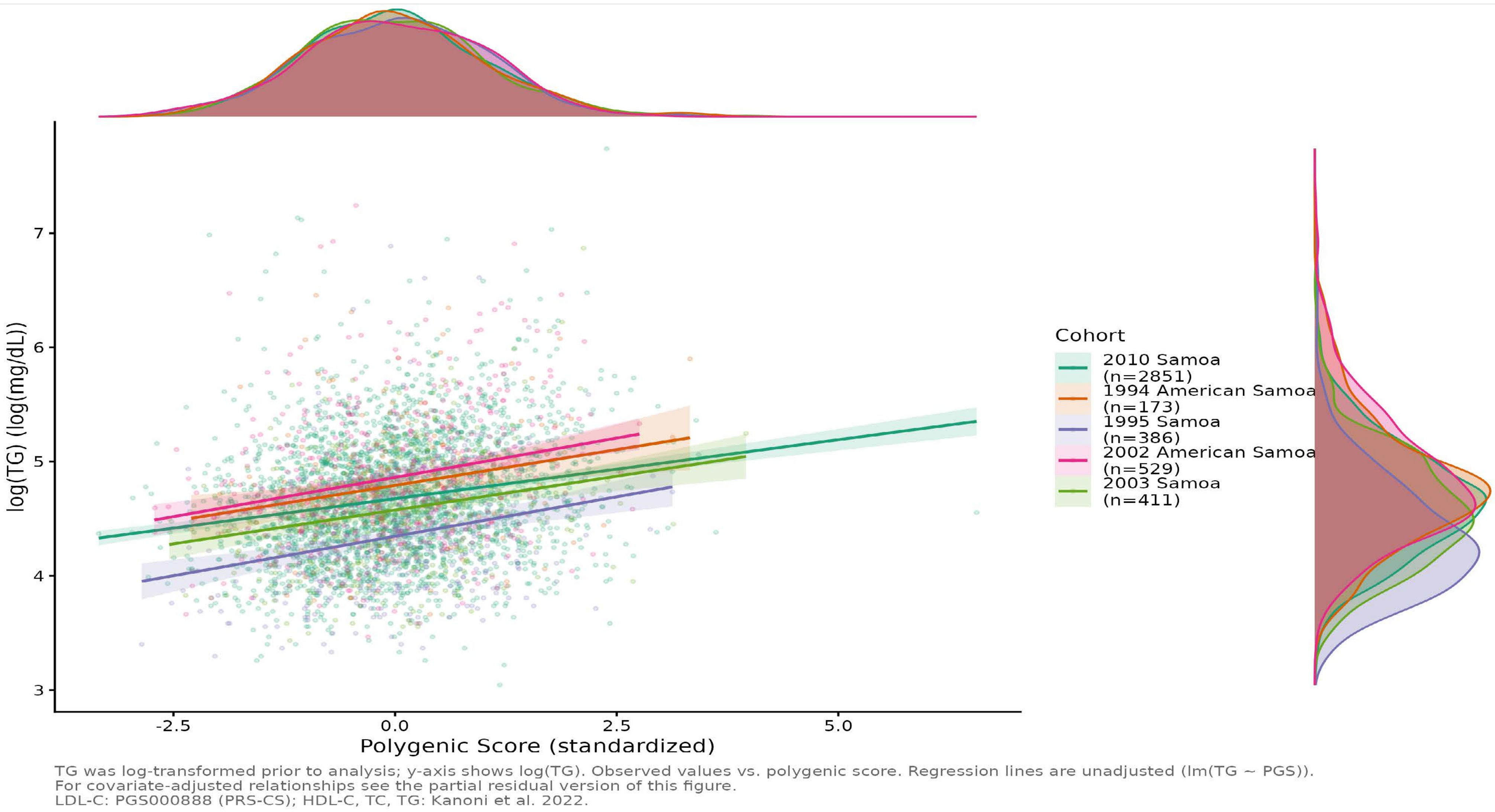


Figure S4. Distribution and polygenic score associations for triglycerides (TG) across five Samoan cohorts. Observed log−transformed TG (TG was natural log−transformed prior to analysis to reduce right skew) vs. standardized polygenic score (PGS), with unadjusted linear regression lines and 95% confidence bands per cohort. Marginal density plots show the distribution of PGS (top) and log−transformed TG (right) by cohort. PGS source: Kanoni et al. (2022).

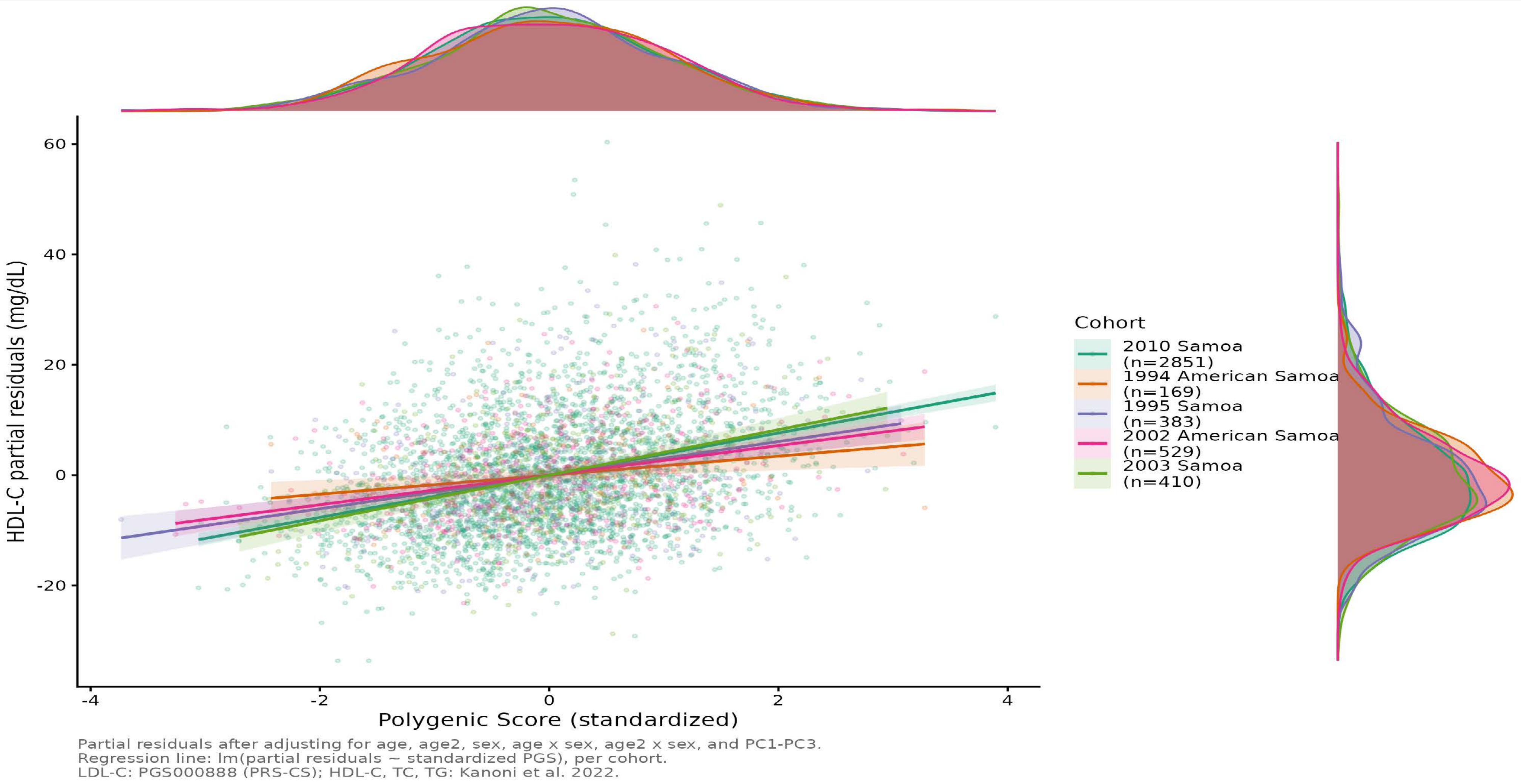


Figure S5. Covariate−adjusted polygenic score associations for HDL−C across five Samoan cohorts. Partial residuals of HDL−C (mg/dL) after adjusting for age, age2, sex, agexsex, age2xsex, and the first three principal components of ancestry (PC1−PC3), plotted against the standardized PGS. Regression lines represent lm(partial residuals ~ PGS), directly corresponding to the covariate−adjusted models reported in the primary analyses. Marginal density plots show the distribution of the standardized PGS (top) and partial residuals (right) by cohort. PGS source: Kanoni et al. (2022).

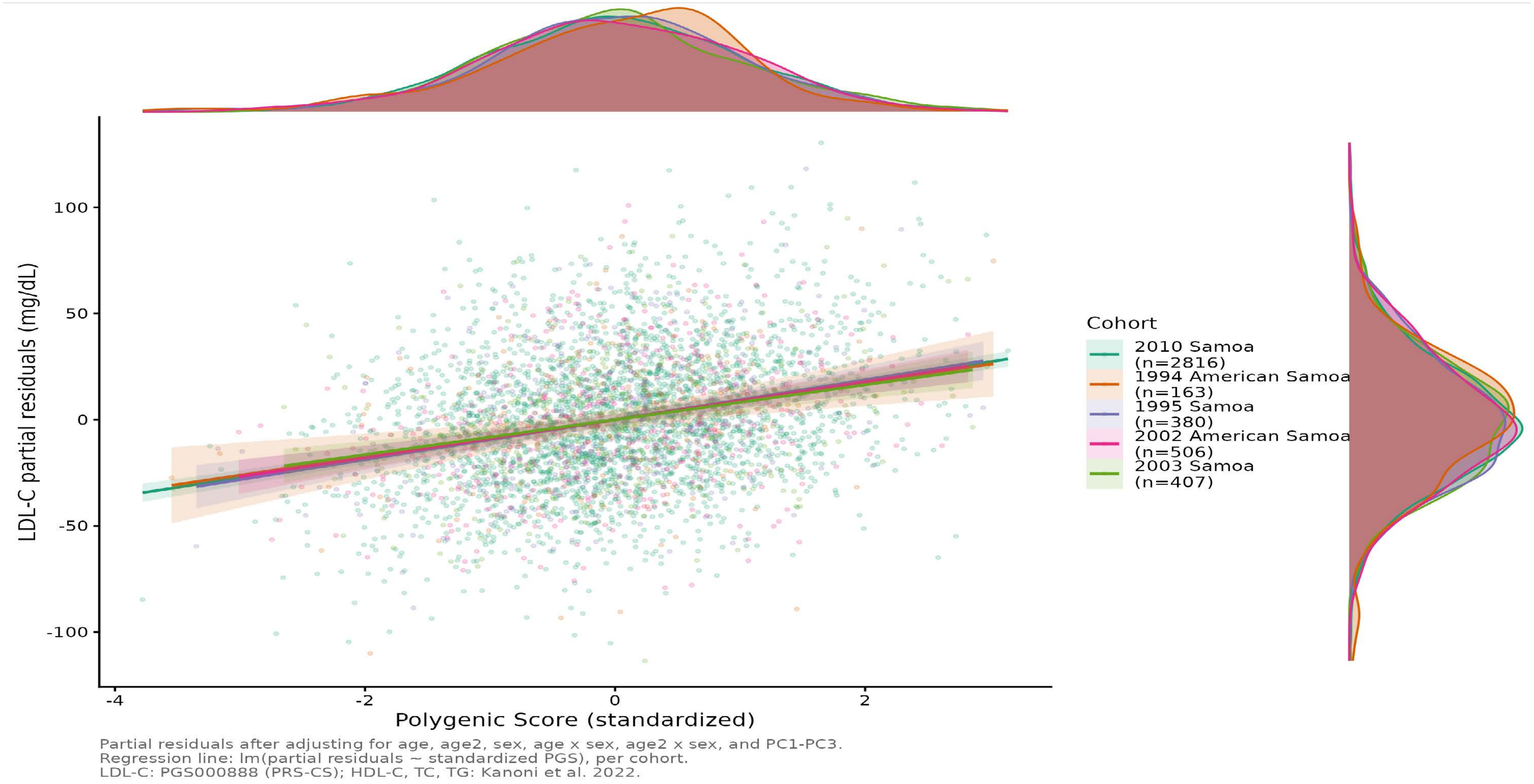


Figure S6. Covariate−adjusted polygenic score associations for LDL−C across five Samoan cohorts. Partial residuals of LDL−C (mg/dL) after adjusting for age, age2, sex, agexsex, age2xsex, and the first three principal components of ancestry (PC1−PC3), plotted against the standardized PGS. Regression lines represent lm(partial residuals ~ PGS), directly corresponding to the covariate−adjusted models reported in the primary analyses. Marginal density plots show the distribution of the standardized PGS (top) and partial residuals (right) by cohort. PGS source: PGS000888, Graham et al. (2021), PRS−CS method.

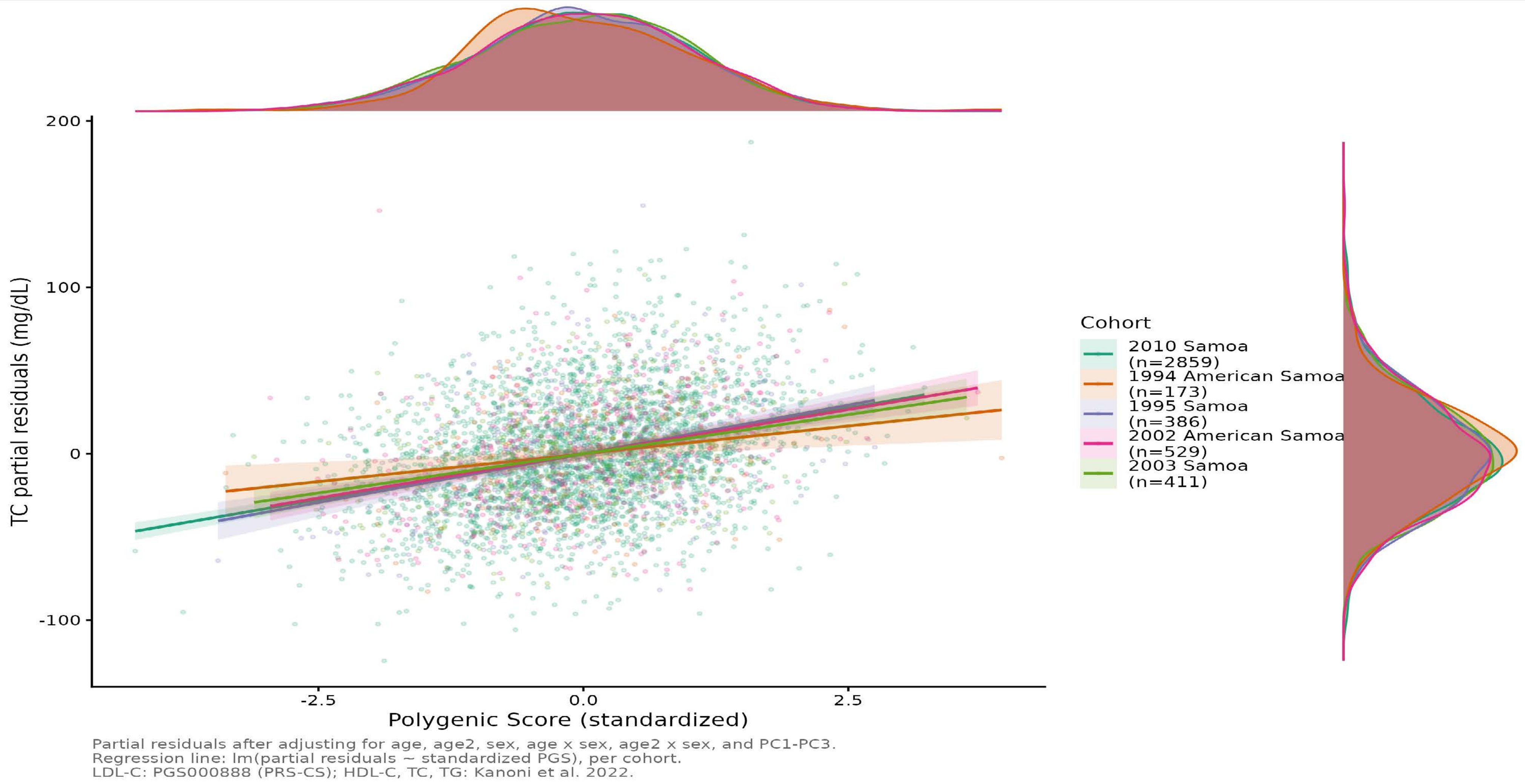


Figure S7. Covariate−adjusted polygenic score associations for total cholesterol (TC) across five Samoan cohorts. Partial residuals of TC (mg/dL) after adjusting for age, age2, sex, agexsex, age2xsex, and the first three principal components of ancestry (PC1−PC3), plotted against the standardized PGS. Regression lines represent lm(partial residuals ~ PGS), directly corresponding to the covariate−adjusted models reported in the primary analyses. Marginal density plots show the distribution of the standardized PGS (top) and partial residuals (right) by cohort. PGS source: Kanoni et al. (2022).

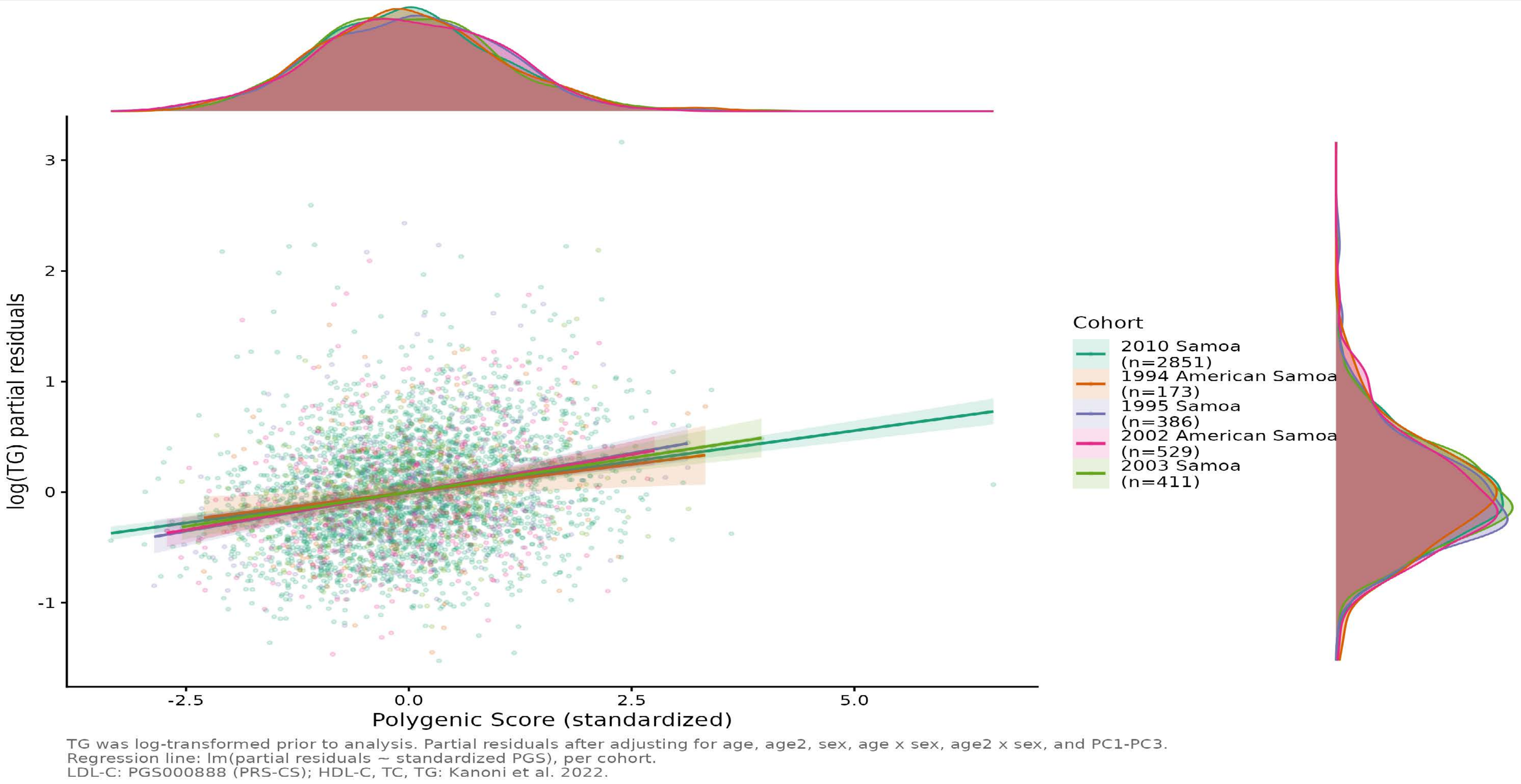


Figure S8. Covariate−adjusted polygenic score associations for triglycerides (TG) across five Samoan cohorts. Partial residuals of log(TG) (TG was natural log−transformed prior to analysis) after adjusting for age, age2, sex, agexsex, age2xsex, and the first three principal components of ancestry (PC1−PC3), plotted against the standardized PGS. Regression lines represent lm(partial residuals ~ PGS), directly corresponding to the covariate−adjusted models reported in the primary analyses. Marginal density plots show the distribution of the standardized PGS (top) and partial residuals (right) by cohort. PGS source: Kanoni et al. (2022).
